# Disparities in Anti-emetic Prophylaxis Care Processes are Predicted by Patient Neighborhood: A Retrospective Cohort and Geospatial Analysis

**DOI:** 10.1101/2024.11.22.24317740

**Authors:** Jiuying Han, Neng Wan, Cameron K. Jacobson, Nathan L. Pace, Cade K. Kartchner, Alexander S. Hohl, Robert B. Schonberger, Douglas A. Colquhoun, Richard P. Dutton, Michael H. Andreae, John F. Pearson

**Affiliations:** School of Environment, Society and Sustainability, University of Utah, Salt Lake City, USA; Department of Anesthesiology, University of Utah School of Medicine, Salt Lake City, USA; Department of Anesthesiology, Yale School of Medicine, New Haven, USA; Department of Anesthesiology, University of Michigan Medical School, Ann Arbor, USA; U.S. Anesthesia Partners, Dallas, USA; Global Change and Sustainability Center, University of Utah, Salt Lake City, USA

**Keywords:** Perioperative Disparities, Antiemetics prophylaxis, Spatial Analysis, Socioeconomic Status, Geography, Electronic Health Registries, Spatial Scan Statistic

## Abstract

**Background:** Social Determinants of Health (SDoH) continue to drive persistent disparities in perioperative care. Our team has previously demonstrated racial and socioeconomic disparities in perioperative processes, notably in the administration of antiemetic prophylaxis, in several large perioperative registries. Given how neighborhoods are socially segregated in the US, we examined geospatial clustering of perioperative antiemetic disparities.

**Methods:** We conducted a retrospective cohort study of anesthetic records from the University of Utah Hospital with 19,477 patients meeting inclusion criteria. We geocoded patient home addresses and combined them with the Census Block Group(CBG) level neighborhood disadvantage (ND), a composite index of from the National Neighborhood Data Archive (NaNDA). We stratified our patients by antiemetic risk score and calculated the number of anti-emetic interventions. We utilized Poisson Spatial Scan Statistics, implemented in SaTScan, to detect geographic clusters of under-treatment.

**Results:** We identified one significant cluster (p < .001) of undertreated perioperative antiemetic prophylaxis cases. The relative risk (RR) of the whole cluster is 1.44, implying that patients within the cluster are 1.44 times more likely to receive fewer antiemetics after controlling for antiemetic risk. Patients from more disadvantaged neighborhoods were more likely to receive below median antiemetic prophylaxis after controlling for risk.

**Conclusions:** To our knowledge, this is the first geospatial cluster analysis of perioperative process disparities; we leveraged innovative geostatistical methods and identified a spatially defined, geographic cluster of patients whose home address census-tract level neighborhood deprivation index predicted disparities in risk adjusted antiemetic prophylaxis.

## 1. Introduction

Social determinants of health (SDoH) continue to drive disconcerting healthcare disparities^1–4^. SDoH are defined as “the societal circumstances in which we are born and grow up, learn and mature, and work and age”: race, ethnicity, education, wealth, insurance coverage, health literacy, etc. impact equitable perioperative processes and outcomes, as fundamental causes of disease^1, 3–5^. Disparities can concern *access* to care, care *processes* and healthcare *outcomes*. We focus on care *processes*, as the means to intervene^2, 3, 6^. To precisely target the underlying causes, to create, test and implement specific countermeasures, we need to define the exact granular mechanisms leading to disparities in *processes* ^4, 5, 7–10^.

### 1.1 Healthcare Disparities in Anesthesiology and Perioperative Medicine

Disparities in care *processes* lead to subpar health *outcomes* in minoritized, marginalized, and migrant populations; differences in care processes based on race and ethnicity or socioeconomic status rather than patient risk factors and co-morbidities, have been a longstanding and troubling aspect of care delivery in the US^1, 3^; In *outpatient* settings, racial and ethnic disparities impact care for diabetes, asthma, and heart disease^11, 12^, as well as veterans care. For *inpatients*, race-based process-of-care surgical disparities^13^ have been described, especially within neonatal and obstetric care^7, 13–15^. Macario et al and our team previously demonstrated process variability in anesthesiology, possibly tied to unconscious bias and negative stereotypes,^2, 5, 16^ as well as disparities in access to chronic pain treatment^10^. We previously demonstrated that individual clinicians provide fewer anti-emetics to people who identify as Black, to those with lower health insurance, and to patients living in zip codes with lower median income^2, 5^. One hypothesis is this arises from clinician prejudice towards their patients^17^: clinicians (sub)consciously detect the social status of their patients in their interaction, and therefore become less likely to diligently elicit for, document, and treat risk factors of postoperative nauseas and vomiting (PONV):^2, 5^ We discussed possible concrete drivers leading to PONV disparities elsewhere.^4^ Given how neighborhoods are socially segregated in the US, the patients home address can provide a proxy for socioeconomic status and race^5, 9^ this motivates our more granular *geospatial* analysis of perioperative process disparities in anesthesia care, to our knowledge the first^9^: Our primary question is whether we can identify clusters of neighborhoods with elevated social deprivation scores whose inhabitants receive less antiemetic prophylaxis after controlling for (PONV) risk^2,18^.

### 1.2 Geospatial Analysis to Investigate Healthcare Disparities

The utilization of geospatial analysis to understand social and environmental influences on perioperative outcomes is a promising emerging science, with studies suggesting zip code may predict health outcomes better than a patients’ DNA^2, 10^. Figure 1 illustrates the various data that can be leveraged for geospatial analysis (e.g. spatial accessibility, air quality, neighborhood deprivation) We discussed the promise and pitfalls of geospatial analysis for health systems and disparity research and the pertinent concepts previously^9^. While more mainstream in public health, very few studies have applied these innovative techniques to perioperative disparities research, especially in conjunction with large scale perioperative registry data, such as Multicenter Perioperative Outcomes Group (MPOG), a leading US perioperative electronic health record registry^18, 19^. The integration with census-tract and block group level social determinants of health derived from publicly available databases (e.g., the National Neighborhood Data Archive (NaNDA)) further augments the power of granular electronic health registries^9^. Not only is geographically-informed health care disparity research ideally suited to studying the impact of patients’ SDoH on their perioperative trajectory, but the information can also be leveraged to provide individual clinicians, teams, and institutions actionable insights on how to improve care processes^4, 7, 9^. While northern Utah and the Salt Lake region have a reputation for homogeneity, the National Equity Atlas ranks Salt Lake County as 208 out of 430 US Counties ranked, with a diversity index of 0.95 (diversity index is a measure of racial/ethnic diversity, with a maximum score of 1.95 if all ethnic groups equally represented), while the US as a whole ranges from of 0.22 to 1.58, making our study population comparable to a broad swath of mid-size US Counties^20^.

Our innovation was to apply a novel geostatistical method (Poisson Spatial Scan statistic)^21^ to identify neighborhood-level clusters of systematic under-treatment after adjusting for PONV risk. As this methodology represents an alternative statistical approach than classical statistics, we provide Online Supplemental Table 1 to aid readers in interpretation of our results. We chose to utilize geospatial clustering, widely used in disease and crime surveillance,^22, 23^ to explore its use of this technique in the service of healthcare equity. We hypothesized that, driven by individual clinicians’ prejudiced clinical decisions,^2, 5, 17^ patients living in disadvantaged neighborhoods, (with higher density of historically under-served minorities, higher indicators of neighborhood deprivation, and socioeconomic stressors), receive fewer perioperative antiemetics after adjusting for antiemetic risk factors, and that such neighborhoods would be found to cluster geographically^2, 18, 24^.

## 2. Materials and Methods

We performed a retrospective cohort analysis of the University of Utah local MPOG electronic health registry (supplemented with University of Utah Health Epic database), enriched with geocoded SDoH for the year 2021^25^. We adhered to the Strengthening the Reporting of Observational Studies in Epidemiology (STROBE) statement and principles with our checklist included in the Online Supplementary Material^26^. This study was approved and considered exempt on February 9, 2023 by the Institutional Review Board at the University of Utah. All authors approved the statistical analysis plan before analyses began. We describe patient demographic characteristics, PONV risk, and perioperative antiemetic prophylaxis in Table 1.

### 2.1 Geocoding Social Determinants of Health

Patient home addresses were obtained from Epic and then geocoded to latitude and longitude with ArcGIS (ESRI, Inc.)^9, 27^ and spatially matched to Census Block Group (CBG) the boundaries of which were obtained from US Census TIGER/Line 2020 CBG boundaries layer. Any incomplete address information was excluded. We extracted “neighborhood disadvantage”(ND), a composite index of neighborhood deprivation, from the National Neighborhood Data Archive (NaNDA), a publicly available database, and used this as our independent variable.^28^ ND is an average of five US Census indicators, including proportion of the following: non-Hispanic Black, female headed families, households with public assistance or on food stamps, income below federal poverty level, and unemployment, all derived from the United States Census American Community Survey 5-year estimates from 2016-2020. ND is an average of these proportions, therefore as this value increases the relative disadvantage or social stress increases.

### 2.2 Inclusion and Exclusion Criteria

Following the MPOG/ASPIRE quality metric PONV05 Inclusion Criteria,^29^ and utilizing information obtained from the Utah local Epic and MPOG instances, we excluded cases with American Society of Anesthesiologists Physical Classification Status (ASA) of 5 or 6, age<18 years, ICU transfers, and cases performed without general anesthesia (including Obstetric, ECT electroconvulsive therapy, and bronchoscopy procedures). We limited our study area further to the Wasatch Front, a densely populated metropolitan area served by the University of Utah hospital in Salt Lake City, UT, as this better enables geospatial clustering analysis and is more generalizable to urban areas in the US. We present in Figure 2, a Quorum Flow Chart, with missing data noted therein.

### 2.4 Risk-Adjusted Antiemetic Prophylaxis

The novel geostatistical method (Poisson Spatial Scan statistic), required a characterization of patients as receiving relatively more(+1), (similar(0), or fewer(−1)) antiemetic interventions compared to their peers with *similar* PONV risk factors, leading us to define a three-level ordinal variable of “risk-adjusted antiemetic prophylaxis”, or RAAP, similar to the MPOG PONV-05 quality metric; RAAP is specified below.

#### Antiemetic Interventions

The response variable was the administration of appropriate numbers of antiemetic prophylactic interventions. We counted up to six antiemetic interventions (0-1, 2, 3, 4, 5, 6+).^4, 24^ These interventions included the number of and class of anti-emetic prophylaxis (1 point per each), and the use of Total Intravenous Anesthesia (TIVA), as the presence of a propofol infusion, as defined in PONV05, a performance metric developed by MPOG.^25^

#### Postoperative nausea and vomiting risk

Risk factors for PONV likely mediate risk adjusted antiemetic prophylaxis; as they could be unevenly distributed among different racially, ethically, and geographically-defined groups; PONV risk factors could confound the analysis of equitable antiemetic administration.^2, 4, 30^ All PONV risk factors were extracted from our local MPOG instance. We categorized the PONV risk score into six ordinal levels (0-1, 2, 3, 4, 5, 6+), following the PONV05 standard of MPOG.^29^ These PONV risks are defined by PONV05 and follow widely accepted guidelines including female sex (assigned at birth), history of PONV or motion sickness, non-smoker, opioid use (either intra-op or post-op), duration of inhalational anesthesia greater than 1 hour, age < 50 years old, and certain select procedures.

#### Characterizing risk-adjusted antiemetic prophylaxis

First, we stratified our patients by antiemetic risk.^4, 24^ Within each stratum of risk, we calculated the median number of antiemetic interventions. We defined RAAP to describe above median, median, and below median *risk-adjusted* antiemetic prophylaxis (RAAP). Finally, we assigned the three levels (below median, median, and above median) RAAP contingent on the number of prophylactic antiemetic interventions received: a **high** RAAP for an above median number, a **median** RAAP for cases with an intermediate number, and a **low** RAAP for cases with a below median number of antiemetic prophylactic interventions compared to other cases in the same PONV risk stratum. With RAAP, we sought to *contrast* different levels of antiemetic prophylaxis *despite* similar PONV risk, i.e. process disparity driven by neighborhood-level social determinants of health.^4^ Hence, RAAP serves to characterize the relative intensity of antiemetic prophylaxis after controlling for risk, (not as a measure of guideline adherences).^4^ This was due to guideline variability and that the use of adherence to guidelines could obscure underlying disparity within our practice, which includes physicians who trained outside of the US, which runs counter to the purpose of the investigation.

### 2.5 Cluster Analysis

We utilized Poisson Spatial Scan Statistic^21, 31^ implemented in SaTScan (http://www.satscan.org/)^32^ to detect the clusters of under-treated perioperative antiemetic prophylaxis cases in the Wasatch Front. A cluster contains a set of neighboring regions which collectively have higher incidence (of low RAAP) rates than expected if the low RAAP cases were evenly distributed across the study area.

We used univariate and multivariate logistic regressions to investigate the characteristics of the CBG in the identified low RAAP cluster. The outcome was a binary variable indicating whether the CBG belongs to the high-rate low RAAP, or not. We tested previously described or suspected social determinants of health as predictors of disparities:^2, 5^ percentage of non-White population, percentage of population with education less than high school, percentage of population below poverty rate, percentage of population married, percentage of elderly (> 65years), and percentage of renters.

## 3. Results

### 3.1 Description of the Population and Quorum Patient Flow Diagram

51,809 anesthetic case records of patients undergoing surgery at the University of Utah were extracted from the Utah MPOG 2021 dataset. After excluding cases with missing data, a final dataset of 19,477 cases were analysed in cluster analysis, as detailed in Figure 2, Quorum Flow Chart, with missing data noted therein. Table 1 lists detailed characteristics of the original cohort as well as the final analysis cohort after applying our exclusions criteria.

The median age in the final cohort was 48 years (IQR 34-63); more than 95% of patients underwent a general anesthetic, often lasting longer than 1 hour (40%); the administration of opioid medication was almost always part of the anesthetic plan (98.5%). ASA class was mostly 1-3 (12%, 47% and 37%, respectively). Approximately one-tenth of patients reported a history of PONV. Most patients were non-smokers (98.6%). The median number of PONV risk factors was 4(IQR 3-4); patients received a mean of 2 (IQR 2-3) prophylactic antiemetic interventions.

The final cohort had approximately the same percentage of white patients (79.5% original vs. 78.2% final, SMD=0.03), similar sex distribution (56% female original vs. 56% final, SMD=0.01, and similar smoking status (99% non-smoking original vs. 99% final, SMD=0.00) while the final cohort had slightly more risk factors (3.46 vs. 3.79, SMD=0.29), interventions (1.88 vs. 2.44, SMD=0.55), and higher prevalence of a lower ASA(ASA1-2 50% original vs. 59% final, SMD=0.27).

### 3.2 Uni- and Bivariate Analysis

In total, we found 2,260 low RAAP cases, 7,358 median RAAP, and 9,589 high RAAP cases in our final dataset. RAAP reflects *risk adjusted antiemetic prophylaxis,* as defined in the methods. The association between socioeconomic neighborhood-level and demographic patient factors and RAAP is presented in a bivariate tabulation (Table 2). Patient self-identified race predicted RAAP. Living in more disadvantaged neighborhoods (e.g., Q4 = neighborhood disadvantage 4th quartile) was associated with receiving fewer antiemetic interventions after adjusting for risk (17% low RAAP); in other words, compared to patients living in more affluent neighborhoods (Q1 = 1st neighborhood disadvantage quartile with 49% high RAAP), clinicians administered fewer antiemetic interventions, even after adjusting for risk, in cases of patients living in census block groups with higher neighborhood disadvantage scores. Additionally, after controlling for PONV risk, patients who self-identified as White received more antiemetic interventions (80% high RAAP, p<0.01), and rarely fewer antiemetic interventions (only 10% low RAAP, p<0.01) while Non-White patients more often received fewer interventions after controlling for risk (15% low RAAP, p<0.01), compared to White patients in similar PONV risk strata. These associations are congruent with the results of our more complex statistical models, described below.

The bivariate analyses showed that all the variables of interest except age were significantly related to being inside the cluster, with the percentage of non-White, percentage of male, percentage of education less than high school, percentage of population below poverty level, and the percentage of renter being positively related to belonging to a high-rate cluster, while the percentage of married population being negatively related to belonging to a cluster.

### 3.3 Geospatial Analysis

We identified one significant cluster (p < .001) of low risk adjusted antiemetic prophylaxis (RAAP) cases in Wasatch Front (Figure 3), comprising 101 census block groups (CBGs). The cluster was located around the geographic centre of Salt Lake City, and included West Valley City, South Salt Lake, and Taylorsville. The cluster had a total of 1,934 included participants residing within it, or approximately 9.93% of the study population. The RR of the whole cluster was 1.44 (p<.001), implying that the risk to receive less risk adjusted antiemetic prophylaxis (low RAAP) was 1.44 times higher for patients living *within* the cluster than for those living in other CBGs along the Wasatch Front. The CBG level RR within the cluster ranged between 0 to 3.73 (with RR>1 being a high relative risk), meaning the highest risk of low RAAP was 3.73 times higher living in CBGs within versus outside the cluster.

After adjusting for all listed covariates, we found that percentage of age greater than 65 years in a CBG cluster became highly significantly associated with a low RAAP cluster, which would represent guidance congruent care as age protects against PONV. However, we similarly demonstrated that the percentage of non-White people in a geographic area was also associated with low RAAP, which represents an unexplained process disparity. The larger the percentage of age greater than 65 and the percentage of non-White, the more likely the CBG belonged to the low RAAP cluster. In addition, the percentage of males was positively related to belonging to a low RAAP cluster, but with borderline statistical significance, (again indicating guidance congruent care, as female gender is positively associated with PONV risk).

## 4. Discussion

In our retrospective cohort of patients at the University of Utah local MPOG electronic health registry,^25^ we leveraged a novel geostatistical method to identify a spatial cluster of patients receiving less risk-adjusted antiemetic prophylaxis (RAAP) than peers in similar PONV risk strata.^32^ We identified one significant cluster (p < .001) of low RAAP cases in Wasatch Front (Figure 3), comprising 101 CBGs.

The cluster identified is an area of low RAAP cases with a high neighborhood disadvantage score and a relatively higher percentage of Hispanic, non-white patients. Low RAAP, defined in the methods, reflects process disparities in risk-adjusted antiemetic prophylaxis. Our findings are consistent with a direct association between neighborhood disadvantaged and risk-adjusted antiemetic prophylaxis. Our results remained statistically significant in multi-variate analysis, where we demonstrated that non-white received lower RAAP than self-identified white participants. Uni- and bivariate tabulation of our data (Table 2) corroborated our principal geospatial analysis, while the association of male gender and older age in census block groups with low RAAP were consistent with guidance congruent care, as both patient characteristics are associated with lower PONV risk.

### 4.1 Interpretation of Principal Findings

Only individual patient-level PONV risk factors should be the guiding principle of risk adjusted antiemetic prophylaxis (RAAP);^2, 4, 5, 24^ neither patient race and ethnicity, nor neighborhood disadvantage should impede equitable risk-adjusted antiemetic prophylaxis.^2, 18, 24^ By identifying geographic clusters of low RAAP, representing populations experiencing disparities in antiemetic prophylaxis after adjusting for risk, we visualized how geographically defined social determinants of health drive healthcare process disparities within Anesthesiology.^9^ Our finding that those within a disadvantaged geographic area, in this case the Hispanic dominated West Valley of Salt Lake County, are over 40% less likely to receive RAAP is troubling, and reinforces the utility of geospatial analysis to identify populations at risk of process disparities. It should be further noted that the use of the Spatial Scan statistic examines individual CBGs in the context of neighboring CBGs: the ability to analyse beyond administrative boundaries can capture broader area characteristics, an advantage when examining sub-divisions within a broader area of disparity.

### 4.2 Significance of Geospatial Process Disparities in the Literature

Why would a patient’s home address determine how many antiemetics they receive, as we demonstrated in this geospatial analysis? Geographic analyses in perioperative medicine have been sparse^33–35^ and prior work tended to focus on patient *outcomes* rather than on *process* disparities. Geographic clusters of disparities in care have been well documented in various areas of medicine, including diabetes, asthma, heart disease, veterans care, surgical care, as well as neonatal care.^7, 11–15^ Our findings also build on process outcomes in paediatric anesthesiology and our own work in obstetric care pain medicine, where cancellation rates were found to be higher in communities with lower socioeconomic status.^10, 19, 34, 36, 37^. Our findings also add to our prior research on disparities in risk-adjusted antiemetic prophylaxis driven by patient- and neighborhood-level social determinants of health (patient race, insurance, and median income in zip code).^2, 5^. Our results are consistent with these prior findings but examine perioperative process disparities through a geospatial lens with granular geostatistical methods, considering proximity statistically.^9, 21^

### 4.3 Promise of Geospatial Analysis of Disparities

The innovation of our study is to perform a geospatial analysis of perioperative process disparities, leveraging equitable risk adjusted antiemetic prophylaxis as a case study.^2, 5^ This represents an important step forward in terms of ability to ameliorate disparities, as geospatial drivers can be re-evaluated over time, e.g. after mitigation measures are put in place to address disparities.^9^ The benefits of exploring process differences by neighborhood was also noted in studies in Scotland and California;^38^ identifying specific neighborhood clusters provided for targeted outreach to improve diabetes care.

Our analysis of local catchment areas with disparities in care *processes*, for which we are individually accountable as clinicians,^5^ can make for a different conversation within a department and among clinicians about potential modifications to practice patterns; these may ultimately drive many outcome disparities which some may have traditionally considered beyond our direct influence and scope of practice.^7^ Outcome differences may be influenced by environmental factors (e.g., pollution) while differences in risk adjusted processes, under the primary domain of the individual anesthesia provider in the operating room, leave fewer excuses.^4^ In this way, a focus on process provides greater insight into clinician behaviours and relationships with patients and opens the door to potential novel avenues to improve care.^4, 10^

Studies from Taiwan and the United Kingdom National Health Service (NHS) system show a complex interplay between care disparities and socioeconomic status of the neighborhood demonstrating that even in systems with near universal healthcare coverage, socioeconomic status can drive disparities in the delivery of care.^39, 40^ In contrast, other studies have suggested that neighborhood characteristics have only a modest impact on some care processes and outcomes,^41^ while the influence of race may overshadow neighborhood characteristics.

### 4.4 Limitations and Potential Bias in our Methods

Our study is not without its limitations. First, limiting generalizability, we confined the study population to the catchment area of the University of Utah Hospital, limited to one Health system and one group of anesthesiologists. However, our prior studies around race have examined multiple hospitals in the large perioperative electronic health registries with analogous disparities identified.^2, 5^ Furthermore, racial and ethnic diversity of populations vary across the US, somewhat limiting the external validity of our study to states with lower minority representation. Low RAAP areas had low socioeconomic status and higher rates of Hispanic populations. This could result in autocorrelation and bias as it is possible that neighborhood SES and geographic factors were less salient than race but could not be fully separated in our model due to our small patient cohort. Expanding our approach to include areas of economic diversity among the Hispanic or other minoritized populations (e.g. Texas, South Florida, California) would further elucidate this relationship. The MPOG registry and our local dataset does not provide data on the race, gender or ethnicity of the *clinicians* assigned to a particular anesthetic case. We were therefore unable to investigate if congruence of identity characteristics between patients and clinician for a given case had an impact on clinician adherence with best antiemetic practices. Furthermore, the assignment of staff to specific clinical locations could introduce bias in our results, especially if a single sub-group regularly practices at a given location; however as the vast majority of cases take place at the main University of Utah clinical campus, the potential for this bias is minimal.

Another limitation, inherent in all geographic epidemiological retrospective studies, is the ecological fallacy;^9, 42^ we did not have individual self-identified race for all patients, but mostly aggregate data at the neighborhood census-block group level. Despite joining some individual data with group-level data, the risk of ecological bias, while mitigated is still present risk. We acknowledge potential *missing data bias*, since we were only able to identify 93.1% of patients’ addresses to the level of the street point address, (which is, however, consistent with other geocoding results).^9, 43^ We also acknowledge that limiting the study area to the Wasatch Front, which was done to ensure contiguous CBGs for geospatial clustering analysis, may have resulted in exclusion of rural disparities. We also did not examine the race of clinicians or performed any analysis by clinician characteristics.^4^ A few outliers could have skewed the cohort, though this is unlikely given the inclusion of over 100 anesthesiologists. More detailed strength and weaknesses of our geospatial analysis approach are discussed in the Online Supplementary Material.

## 5. Conclusions

Demonstrating the utility of a novel geostatistical method in a retrospective cohort of anesthesia case records from the local MPOG electronic health registry at the University of Utah, we identified a spatial cluster of patients in the West Valley area of Salt Lake City, UT, receiving less risk-adjusted antiemetic prophylaxis (Figure 3); they also represented a population from CBGs with relatively high neighborhood deprivation. Our results are consistent with prior work that demonstrated disparities in antiemetic prophylaxis based on *patient-*level factors (e.g., race),^2^ but adding a *neighborhood*-level, spatial dimension to perioperative healthcare disparities and health systems research, with many potential applications to elucidate geographic, social, and environmental drivers of health care process and outcome (Figure 1).^9^

## Data Availability

All data produced in the present study are available upon reasonable request to the authors.

## 6. Authors Contributions

All listed authors meet the criteria for authorship based on the recommendations of the International Committee of Medical Journal Editors (ICMJE). Further detail on each author’s contribution is as follows:

Jiuying Han: Played a pivotal role in conceptualizing and designing the study. Led the software development, formal analysis, investigation, and data curation. Primarily responsible for writing the original draft, and significantly contributed to the review and editing process. Provided visualization elements. Gave final approval of the version to be published and agrees to be accountable for all aspects of the work.

Neng Wan: Key contributor in conceptualizing the study and shaping its methodology. Provided resources, administered the project, and was involved in funding acquisition. Engaged in the writing, review and editing of the manuscript. Gave final approval of the manuscript and agrees to be accountable for all aspects of the work.

Cameron Jacobson: Contributed significantly to the study’s methodology and software development. Actively involved in the investigation, acquisition and data curation processes. Participated in the drafting, review and editing of the manuscript. Approved the final version to be published and agrees to be accountable for all aspects of the work.

Michael Andreae: Central in conceptualizing the study and developing its methodology. Provided resources, supervised the project, and administered the project’s execution. Acquired funding. Co-Authored the original draft and engaged in the manuscript’s review and editing. Approved the final manuscript for publication and agrees to be accountable for the work’s integrity.

Nathan Pace: Involved in conceptualizing the study and shaping its methodology. Contributed resources, and contributed to analysis of data. Played a significant role in reviewing and editing the manuscript. Approved the final manuscript and agrees to be accountable for all aspects of the work.

Cade Kartchner: Involved in study conceptualization, software development, investigation, and data acquisition and curation. Actively contributed to the writing, review and editing of the manuscript. Approved the final manuscript and agrees to be accountable for all aspects of the work.

Alexander Hohl: Played a significant role in conceptualization of study and developing the study’s methodology. Active role in interpretation of data. Involved in the manuscript’s drafting, review and editing, as well as its validation. Approved the final manuscript and agrees to be accountable for all aspects of the work.

Robert B. Schonberger: Involved in the conceptualization of the project and contributed to the manuscript’s drafting, review and editing, and validation. Active role in interpretation of data. Approved the final version to be published and agrees to be accountable for all aspects of the work.

Douglas A. Colquhoun: Involved in interpretation of the study analysis, reviewing and editing the manuscript, as well as its validation and conception of study design. Gave final approval of the manuscript and agrees to be accountable for all aspects of the work.

Richard Dutton: Actively involved in the project conceptualization, as well as manuscript’s drafting, review and editing, and validation. Approved the final manuscript and agrees to be accountable for all aspects of the work.

John Pearson: Central in conceptualizing the study and developing its methodology. Provided resources, supervised the project, and was involved in funding acquisition. Co-authored the original draft and significantly contributed to the manuscript’s review and editing. Approved the final manuscript for publication and agrees to be accountable for the work’s integrity.

## 7. Acknowledgements

The authors would like to acknowledge Beca Chacin of the University of Utah Department of Anesthesiology for her contribution to creating Figure 1.

## 10. Appendices

Supplemental Table 1 describes the differences in methodological approach and interpretation between classical statistics typically seen in the biomedical literature, and our geospatial analysis approach more typically presented in epidemiology and geography. Of the key differences, it is worth noting that statistical significance is interpreted in classical statistics with p values and ORs, among other tests, while the salient findings in clustering techniques involve visual interpretation and multiple testing of clusters. Another key takeaway is that classical statistics builds no spatial association between datapoints, therefore it does not take spatial proximity into account. This contrasts with geographical approaches that utilize spatial information in order to perform tests, with proximity of data points and values being a key consideration.

## 11. Declaration of generative AI and AI-assisted technologies in the writing process

During the preparation of this work the authors used ChatGPT 4, from OpenAI, in order to edit manuscript for clarity of language and to ensure adherence to the STROBE checklist. After using this tool/service, the authors reviewed and edited the content as needed and take full responsibility for the content of the publication.

## 13. Tables

**Table 1:**
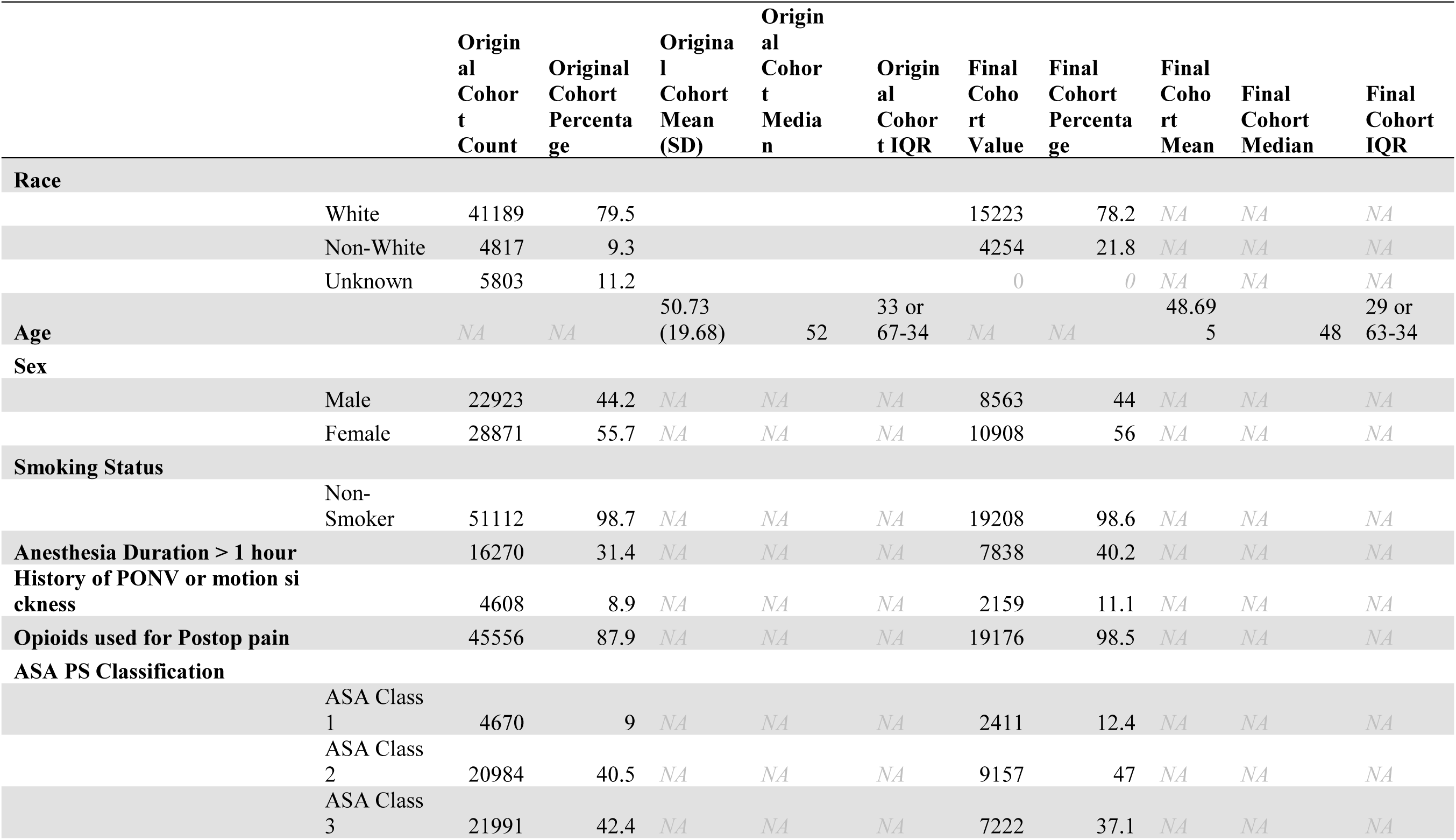
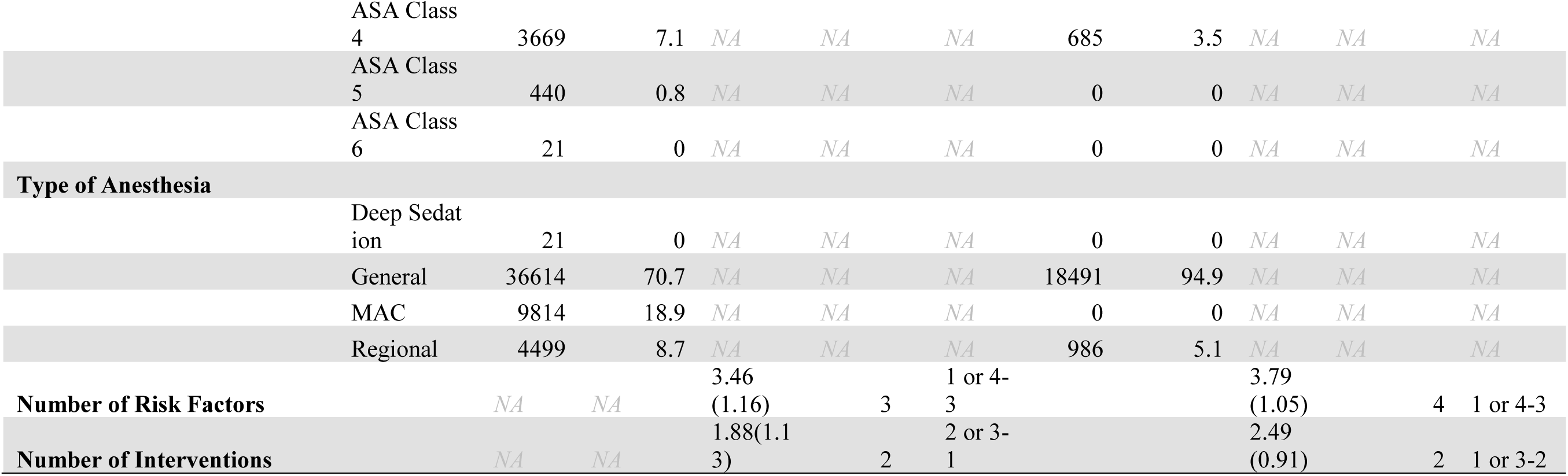
Description of initial and final cohort.

Table 1 details characteristics of our population prior and after applying exclusion criteria and filtering of data to the Wasatch Front for the purpose of our geographic analysis. Characteristics of the final study set with home addresses in the target area are listed in the Final Cohort Columns.

Most patients in the final cohort had a median age of 48 (with an interquartile range of 29) and received a general anesthetic of more than 1 hour duration, including the administration of opioid medication. Their risk classification according to the American Society of Anesthesiology was mostly 1-3. Only ten percent reported a history of PONV, and most were non-smokers. For a median of four PONV risk factors, they received on average about 2 prophylactic antiemetic interventions.

The focus on the Wasatch front area shrunk the total number of patients from 51,809) to 19,477 in the final cohort analysed, and increased the relative proportion of Non-White patients, (typically clustered in urban areas in Utah); the prevalence of risk factors and the number of interventions also changed. Percentages have been rounded to the nearest tenth place with some factors being described by the Mean, Median, and Interquartile Range (IQR) of the data represented.

**Table 2:** Risk-Adjusted Antiemetic prophylaxis and neighborhood disadvantage.

| **Category** | **High RAAP** | **Median RAAP** | **Low RAAP** | **Total** |
| --- | --- | --- | --- | --- |
| **Q1** | 2390 (49.1%) | 1788 (36.7%) | 692 (14.2%) | 4870 |
| **Q2** | 2331 (47.9%) | 1858 (38.2%) | 680 (14%) | 4869 |
| **Q3** | 2128 (43.7%) | 1962 (40.3%) | 779 (16%) | 4869 |
| **Q4** | 2001 (41.1%) | 2033 (41.8%) | 835 (17.1%) | 4869 |
| **Total(Q1-Q4)** | 8850 (45.4%) | 7641 (39.2%) | 2986 (15.3%) | 19477 |
| **White** | 7097 (80.2%) | 820 (9.3%) | 933 (10.5%)* | 8850 |
| **Non-White** | 5990 (78.4%) | 754 (9.9%) | 897 (11.7%)** | 7641 |
| **Unknown** | 2136 (71.5%) | 380 (12.7%) | 470 (15.7%)** | 2986 |
| **Total (Race)** | 15223 (78.2%) | 1954 (10%) | 2300 (11.8%) | 19477 |

Table 2 demonstrates how home address and Race predicted risk-adjusted antiemetic prophylaxis in a descriptive, bivariate comparison. Populations living in more disadvantaged neighborhoods (e.g., Q4 = neighborhood disadvantage 4^th^ quartile) on average received fewer antiemetic interventions after adjusting for risk 17.1% low RAAP), hence they were relatively undertreated compared to patients living in more affluent neighborhoods (Q1 = 1^st^ neighborhood disadvantage quartile with 49% high RAAP). Additionally, people who self-identified as White mostly received many antiemetic interventions after controlling for risk (only 10% low RAAP) compared unknown patients who more often received fewer interventions after controlling for risk (15% low RAAP,). Levels of RAAP are tabulated in three columns, (RAAP is defined in the methods section: below median, median, and above median RAAP reflected disparities in risk-adjusted antiemetic prophylaxis). The Neighborhood Disadvantage (ND) scores in the rows, (explained in the method section as well), were separated into quartiles with the first quartile Q1 being the lowest score (meaning the least disadvantaged) and the fourth quartile Q4 being the highest disadvantaged score (meaning the most disadvantaged).

Finally, an asymptotic generalized Pearson chi-squared test was performed to determine the statistical significance between race categories (white, non-white, etc) and RAAP. This was performed to compare White: Non-white(* p=0.003), White: Unknown (**p<0.001) and Non-white: Unknown (**p<0.001).

## 14. Figures

**Figure 1:**
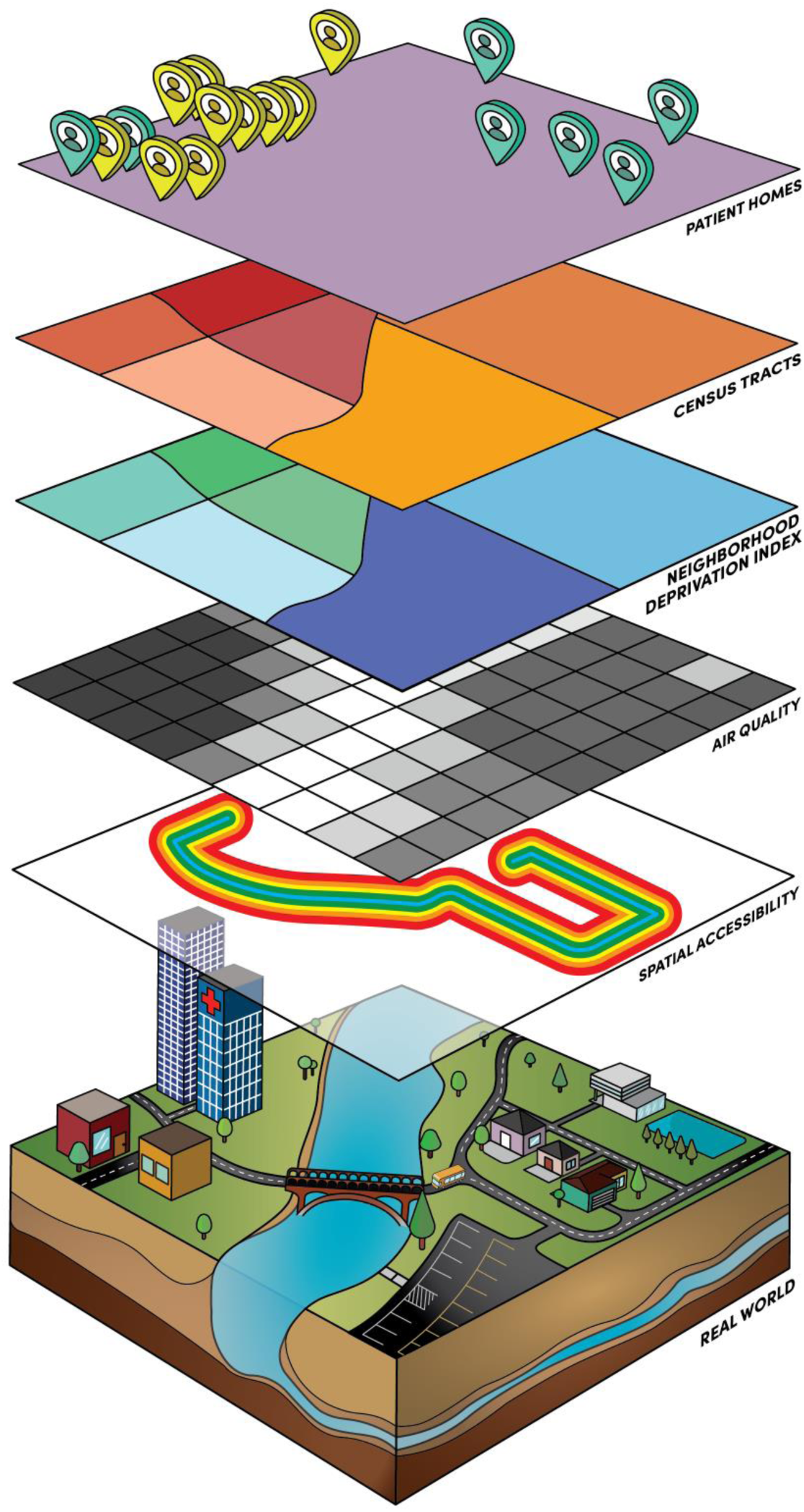
Geographic Analysis and Social Determinants of Health.

Figure 1 demonstrates the ability of Geographic Information Systems (GIS) to integrate various sources of data into a comprehensive model of patient exposures and social determinants of health (SDOH).^1, 9^ The bottom layer depicts the real world. The top layer presents geocoded patient home addresses. The Census-tract (CT) layer can be associated with a wide variety of demographics. Next, the neighborhood deprivation index presents a composite of CT data. Air quality in the layer below differs from others in that everyone is exposed. Finally, spatial accessibility focuses on public transportation, analysing transit stop locations and walking distance.

Geospatial analysis refines healthcare disparity research and helps elucidate the relative contribution of race, economic and geographic-based health disparities. The progression of diabetic foot ulcers to amputation exemplifies this: Researchers demonstrated disparities in care processes in areas of high neighborhood deprivation both in California and Scotland.^34^ Patients in lower socioeconomic neighborhoods had worse care and outcomes due to a lack of preventative care from socioeconomic factors leading to amputation, whereas better access could have prevented this.^34^ The use of neighborhood-level geographic factors in perioperative research has so far been limited and often requires geocoding of patients addresses to extract additional SDoH.^9, 29^

**Figure 2:**
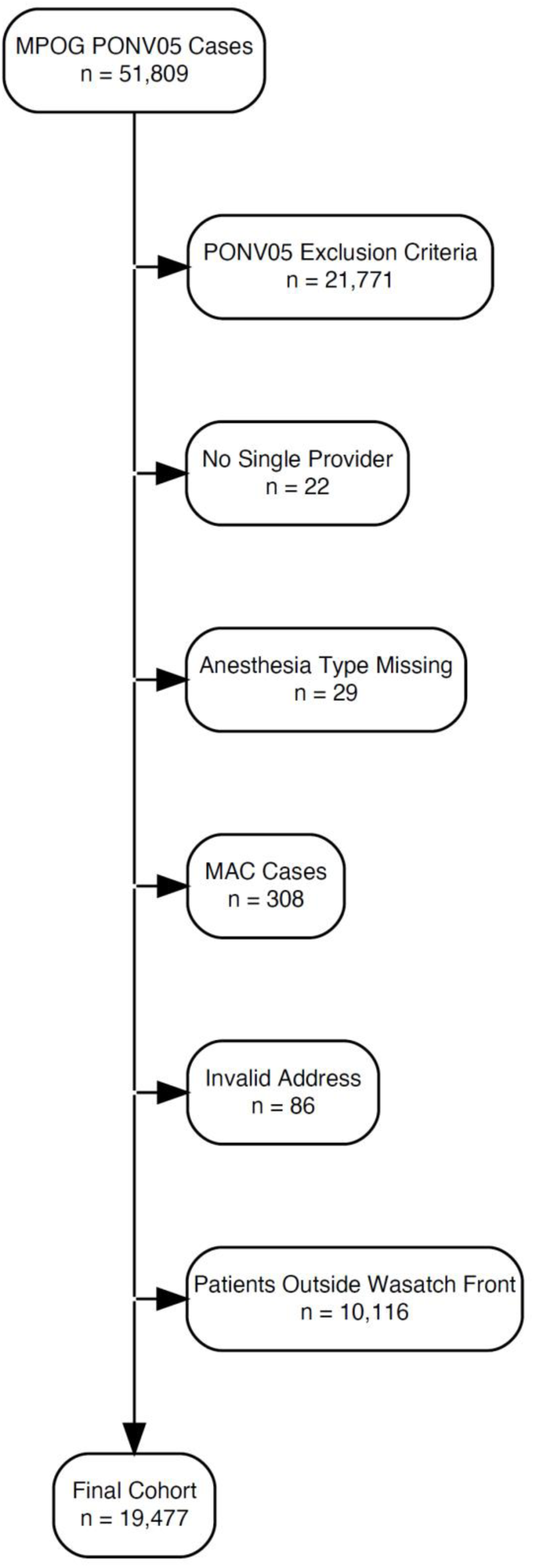
Quorum Patient Flow Diagram.

The initial cohort (n=51,809) included patients who underwent surgery at the University of Utah Hospital during 2021. Data was sourced from the PONV05 quality improvement metric at the Multicenter Perioperative Outcomes Group (MPOG) repository. The cohort was further filtered as follows: Missing or Invalid Addresses (n=1,705), PONV exclusion criteria (n=20,153), cases where a single anesthesia provider could not be identified (n=22), cases whose anesthesia type was MAC (n=335), and finally, patients who lived outside the Wasatch Front (n=10,117). The final cohort included 19,477 cases.

**Figure 3:**
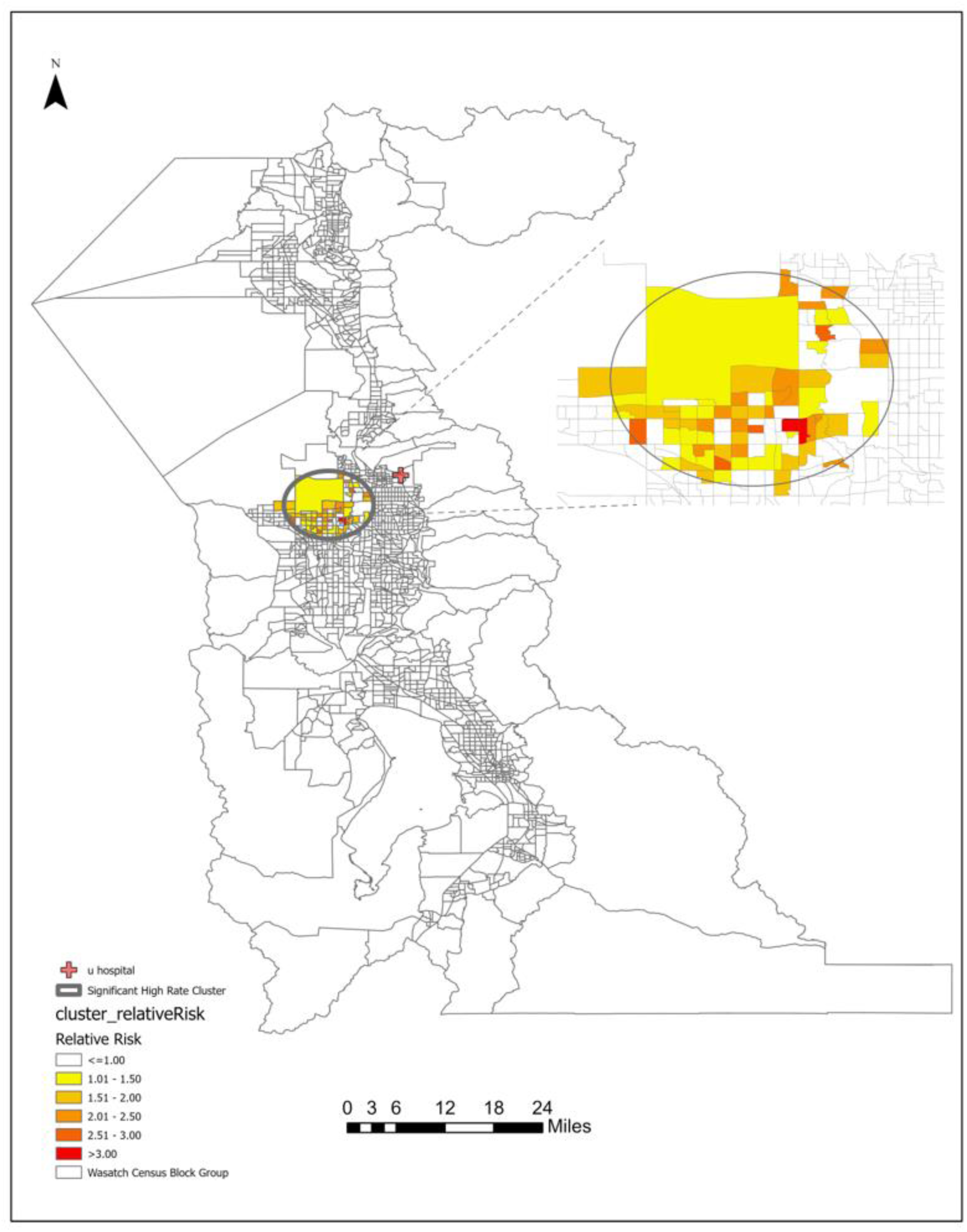
Geospatial Clustering Results.

This map represents the study area of the Wasatch Front, with Census Block Groups outlined in grey. The legend indicates the relative risk of under-treatment from least to greatest as a result of the Satscan methodology.^19^ As noted, the colours indicate only where a contiguous and statistically significant cluster of any type of process disparity in risk adjusted antiemetic prophylaxis (below median, median, versus above median RAAP) was found. In our analysis, only an area of under-treatment was identified, and it corresponded to a region of Salt Lake County with a high prevalence Hispanic population.

