## Supplemental Table for "Disparities in Anti-emetic Prophylaxis Care Processes are Predicted by Patient Neighborhood: A Retrospective Cohort and Geospatial Analysis"

**Supplementary Material**

**Strength and Weaknesses of Different Spatial Analysis Approaches**

| **Aspect** | **Classical Regression Modeling** | **Geospatial Clustering** |
| --- | --- | --- |
| **Unit of Analysis** | Individuals, e.g. case, patient | Geographic Unit, e.g. census block group, tract, region. |
| **Statistical Association** | Determination of association between predictor and outcomes | Spatial proximity and influence of clustering on distribution of data points |
| **Emphasis** | Focuses on the relationship between variables regardless of their spatial arrangement | Emphasizes the geographical distribution and spatial arrangement of data |
| **Objective** | Use of hypothesis testing to determine how variables influence outcomes | Geographic focus: identify and analyze spatial clusters, patterns and trends |
| **Methodological Approach** | Linear & Logistic Regression, Bayesian Modeling | Spatial autocorrelation techniques, k-means clustering, SATSCAN. |
| **Data Requirements** | Requires large datasets for robust statistical analysis, sensitive to outliers | Relies on geocoded data, spatial contiguity, and geographical context |
| **Interpretation & Presentation** | Statistical significance and confidence intervals, Odds Ratio | Results interpreted graphically and spatially, with test to determine significant clusters |
| **Management of Spatial Data** | Not emphasis on accounting for spatial relationships, such as dependency or heterogeneity | Approaches and models incorporate spatial dependencies and patterns |

The application of Poisson Spatial Scan statistics in our study offers distinct advantages over numerous alternative clustering techniques.(1, 2) Notably, it corrects for multiple comparisons, adjusts for heterogeneity within the study population, accommodates covariate adjustments, and mitigates pre-selection bias. In comparison to more visually oriented clustering analyses such as Moran's I, Poisson Spatial Scan statistics exhibit superiority by controlling for covariates. Other prevalent methods for cluster detection encompass local indices of spatial association (LISA) and Getis and Ord's local Gi*(d) statistic. LISA calculates the correlation of incidence rates among neighboring regions and identifies clusters based on significant correlations, while Gi*(d) compares rates to the global mean.

However, these two methods define clusters differently from spatial scan statistics, which employ simulation for cluster detection. LISA will be biased when significant variation of population at risk exist among areas, and Gi*(d) is likely to miss less prominent clusters(3, 4). In contrast, spatial scan statistics compare the dataset with a random process, a technique more broadly employed in public health. One potential weakness of Satscan is that it is based on circular scans and therefore may struggle with clusters of other shapes, which is an area that LISA and Gi*(d) excels with. A previous study(5) discovered that while LISA and Gi*(d) can detect the core of a cluster, they may overlook potential peripheral elements. On the other hand, spatial scan statistics better detects the extent of clusters. Additionally, the potential for false-positives using LISA and Gi*(d) further justifies the use of Poisson Spatial Scan statistics in our study.
